# Plasma Metabolomic Profiling in Patients with Rheumatoid Arthritis Identifies Biochemical Features Predictive of Quantitative Disease Activity

**DOI:** 10.1101/2020.09.13.20193664

**Authors:** Benjamin Hur, Vinod K. Gupta, Harvey Huang, Kerry A. Wright, Kenneth J. Warrington, Veena Taneja, John M. Davis, Jaeyun Sung

**Author notes:** Contributed equally as co-senior authors (John M. Davis, Jaeyun Sung).

## Abstract

**Background:** Rheumatoid arthritis (RA) is a chronic, autoimmune disorder characterized by joint inflammation and pain. In patients with RA, metabolomic approaches, i.e., high-throughput profiling of small-molecule metabolites, on plasma or serum has thus far enabled the discovery of biomarkers for clinical subgroups, risk factors, and predictors of treatment response. Despite these recent advancements, the identification of blood metabolites that reflect quantitative disease activity remains an important challenge in precision medicine for RA. Herein, we use global plasma metabolomic profiling analyses to detect metabolites associated with, and predictive of, quantitative disease activity in patients with RA.

**Methods:** Ultra-high performance liquid chromatography-tandem mass spectrometry (UPLC-MS/MS) was performed on a discovery cohort consisting of 128 plasma samples from 64 RA patients, and on a validation cohort of 12 samples from 12 patients. The resulting metabolomic profiles were analyzed with two different strategies to find metabolites associated with RA disease activity defined by the Disease Activity Score-28 using C-reactive protein (DAS28-CRP). More specifically, mixed-effects regression models were used to identify metabolites differentially abundant between two disease activity groups (‘lower’, DAS28-CRP ≤ 3.2; and ‘higher’, DAS28-CRP > 3.2); and to identify metabolites significantly associated with DAS28-CRP scores. A generalized linear model (GLM) was then constructed for estimating DAS28-CRP using plasma metabolite abundances. Finally, for associating metabolites with CRP (an indicator of inflammation), metabolites differentially abundant between two patient groups (‘low-CRP’, CRP ≤ 3.0 mg/L; ‘high-CRP’, CRP > 3.0 mg/L) were investigated.

**Results:** We identified 33 metabolites differentially abundant between lower and higher disease activity groups (*P* < 0.05). Additionally, we identified 51 metabolites associated with DAS28-CRP (*P* < 0.05). A GLM based upon these 51 metabolites resulted in higher prediction accuracy (mean absolute error [MAE]±SD: 1.51±1.77) compared to a GLM without feature selection (MAE±SD: 2.02±2.21). The predictive value of this feature set was further demonstrated on a validation cohort of twelve plasma samples, wherein we observed a stronger correlation between predicted vs. actual DAS28-CRP (with feature selection: Spearman’s *ρ* = 0.69, 95% CI: [0.18, 0.90]; without feature selection: Spearman’s *ρ* = 0.18, 95% CI: [-0.44, 0.68]). Lastly, among all identified metabolites, the abundances of eight were significantly associated with CRP patient groups while controlling for potential confounders (*P* < 0.05).

**Conclusions:** We demonstrate for the first time the prediction of quantitative disease activity in RA using plasma metabolomes. The metabolites identified herein provide insight into circulating pro-/anti-inflammatory metabolic signatures that reflect disease activity and inflammatory status in RA patients.

## Background

Rheumatoid arthritis (RA) is a chronic, autoimmune inflammatory disease primarily affecting the small diarthrodial joints and other organ systems [1-4] that can eventually lead to bone/cartilage erosion, joint deformity, loss in mobility, and organ damage [5]. Known to be associated with a variety of factors, such as genetic susceptibility [6], age [7], sex [8], smoking status [9], and dietary habits [10], RA is diagnosed in nearly 5 per 1000 adults worldwide, and women are 2 to 3 times more likely to develop RA than men [5]. In addition, as is the case with many complex and progressive disorders, patients with RA exhibit vast heterogeneity in clinical symptoms (e.g., joint inflammation, swelling, pain, stiffness) [11], and in responses to methotrexate and other disease-modifying anti-rheumatic drugs (DMARDs) [12]. Furthermore, immune cells (mainly, B-cells, T-cells, and macrophages) and cytokines are known to be implicated in RA pathogenesis [13]. For example, Haringman *et al*. observed that the abundance of macrophages in synovial tissue was positively correlated with disease activity [14], and Chung *et al*. identified significant differences in levels of multiple cytokines (e.g., IL-6, IL-11, LIF) between RA and healthy controls [15]. In this regard, further understanding of the pathophysiological mechanisms that drive either progression or remission in RA disease activity would be important for identifying prognostic factors and developing more effective treatments [5, 16].

Having practical measures of disease activity is essential for determining the course of RA treatment and for monitoring patient response [3]. To this end, several studies have suggested strategies to quantify (or categorize) RA disease activity by using clinical and inflammatory core components, which include, but are not limited to, the number of tender and swollen joints, erythrocyte sedimentation rate (ESR), serum C-reactive protein (CRP) levels, and patients’ pain levels [17-20]. Among these various strategies, the modified Disease Activity Score that considers 28 joints (DAS28) with either ESR (DAS28-ESR) or CRP (DAS28-CRP) is currently one of the most well-recognized and recommended measures in RA [20].

An emerging area of RA research is in using high-throughput metabolomic profiling approaches, which comprehensively measure all small-molecule biochemicals in a biological specimen (e.g., plasma, serum, urine, synovial fluid, etc.) to enable biomarker discovery and novel insights into the biochemical processes governing disease pathophysiology [11, 21-23]. In particular, recent studies have demonstrated the promise of using such metabolomic technologies on patient-derived biospecimens for classifying patients with RA according to their disease activity categories [21, 24, 25], and for identifying metabolic signatures predictive of treatment response [26-29]. For instance, Teitsma *et al*. used metabolomic profiling in serum samples from early RA patients to identify metabolites and metabolic pathways that were significantly associated with sustained, drug-free remission (DAS28 < 2.6) after tocilizumab- or methotrexate-based therapy [24]. Likewise, Sasaki *et al*. identified 15 and 20 metabolites in plasma and urine, respectively, that were differentially abundant between active RA (DAS28-ESR ≥ 3.2) and inactive RA (DAS28-ESR < 3.2) [25]. These findings suggest that a wider application of global metabolomic profiling—coupled with advanced analytics [30]—can lead to the discovery of novel and predictive biomarkers that complement current standard laboratory tests for assessing disease activity in RA.

To date, a global metabolomic profiling analysis to demonstrate the predictive value of blood biochemicals in estimating disease activity scores for patients with RA, has remained elusive. In this study, on 128 plasma metabolomic profiles from 64 RA patients, we utilize a multi-approach analysis to uncover metabolites that reflect and predict RA disease activity. First, we identify metabolites that stratify patients of ‘higher’ (DAS28-CRP ≥ 3.2) and ‘lower’ (DAS28-CRP < 3.2) disease activity groups. Next, we pinpoint specific metabolites that significantly associate with DAS28-CRP. Interestingly, a few of the metabolites identified through these two approaches were able to differentiate between two groups of patients divided according to their C-reactive protein (CRP) levels in blood (‘high-CRP’, CRP > 3.0 mg/L; ‘low-CRP’, CRP ≤ 3.0 mg/L); these metabolites may possibly reflect metabolic perturbations affected by worsening inflammatory activity. Finally, we utilize a machine-learning technique to predict DAS28-CRP with plasma metabolite abundances. Importantly, we find that the feature selection step led to improved performance in predicting quantitative disease activity, and this translated reasonably well to a validation cohort. Taken together, our findings described herein support a key role for high-throughput metabolomic technologies in identifying blood-borne metabolic signatures of RA disease activity, and lay the groundwork for monitoring disease progression and systemic inflammation using blood samples alone.

## Materials and Methods

### Study Population, Subject Enrollment, Sample Collection, and Demographic Characteristics

The study population consisted of consecutive patients with RA attending the outpatient practice of the Division of Rheumatology at Mayo Clinic in Rochester, Minnesota. Eligibility required patients to be adults 18 years of age or older with a clinical diagnosis of RA by a rheumatologist, fulfilling the American College of Rheumatology/European League Against Rheumatism 2010 revised classification criteria for RA [2]. Patients were excluded if they did not comprehend English, were unable to provide written informed consent, or were members of a vulnerable population (e.g., incarcerated subjects). This led to a total of 76 patients fulfilling the eligibility criteria, who were partitioned into two groups (**Table 1**): for the discovery cohort of this study, 64 patients with available blood samples from at least two outpatient visits 6–12 months apart were included (128 total samples); for the validation cohort, 12 patients whose blood samples were available from only a single outpatient visit were included (12 total samples). Demographic and clinical data, including the numbers of tender and swollen joints, patient and evaluator global assessments, CRP (mg/L), body mass index (BMI, kg/m^2^), smoking status, and results for rheumatoid factor (RF, IU/mL) and anti-cyclic citrullinated peptide antibodies (anti-CCP), were collected from the electronic medical records. The patient samples (140 in total) in the study had established disease with mean age 63.54 (range: 32–86), and 69.7% (53 of 76) were female. Disease activity varied from remission to high disease activity, with a DAS28-CRP mean of 3.0 (range: 1.2–7.0). See **Additional file 1** for distribution of DAS28-CRPs corresponding to all study participants.

**Table 1.**
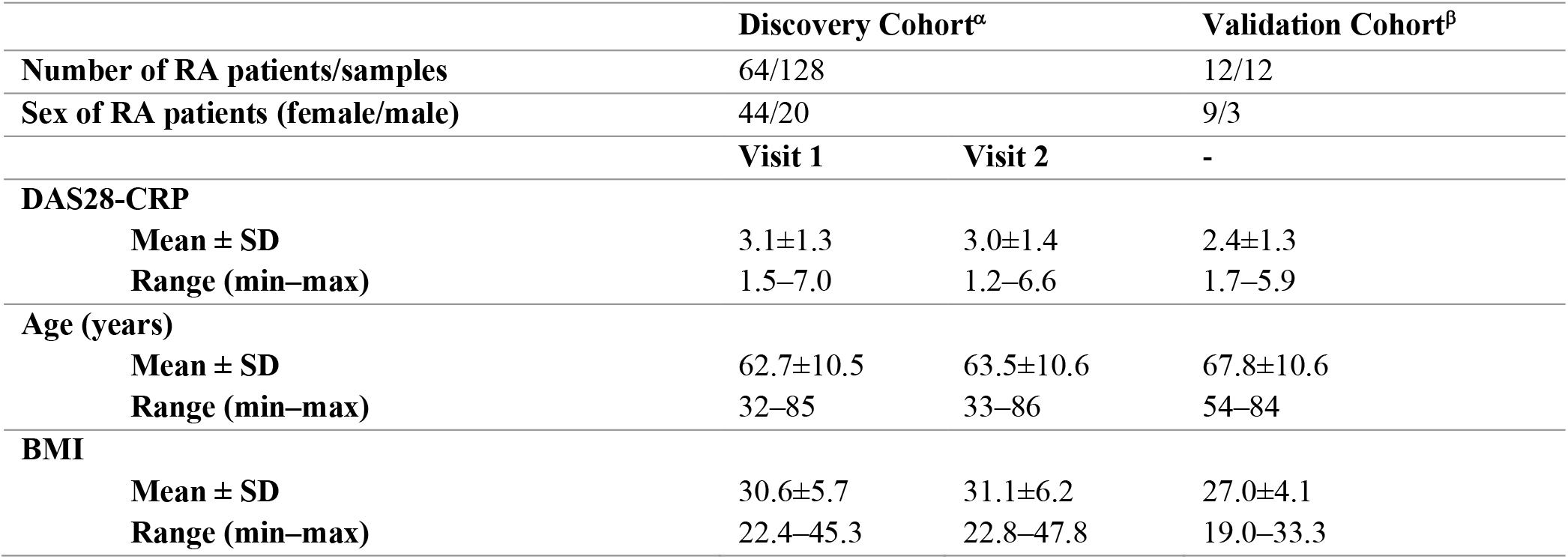
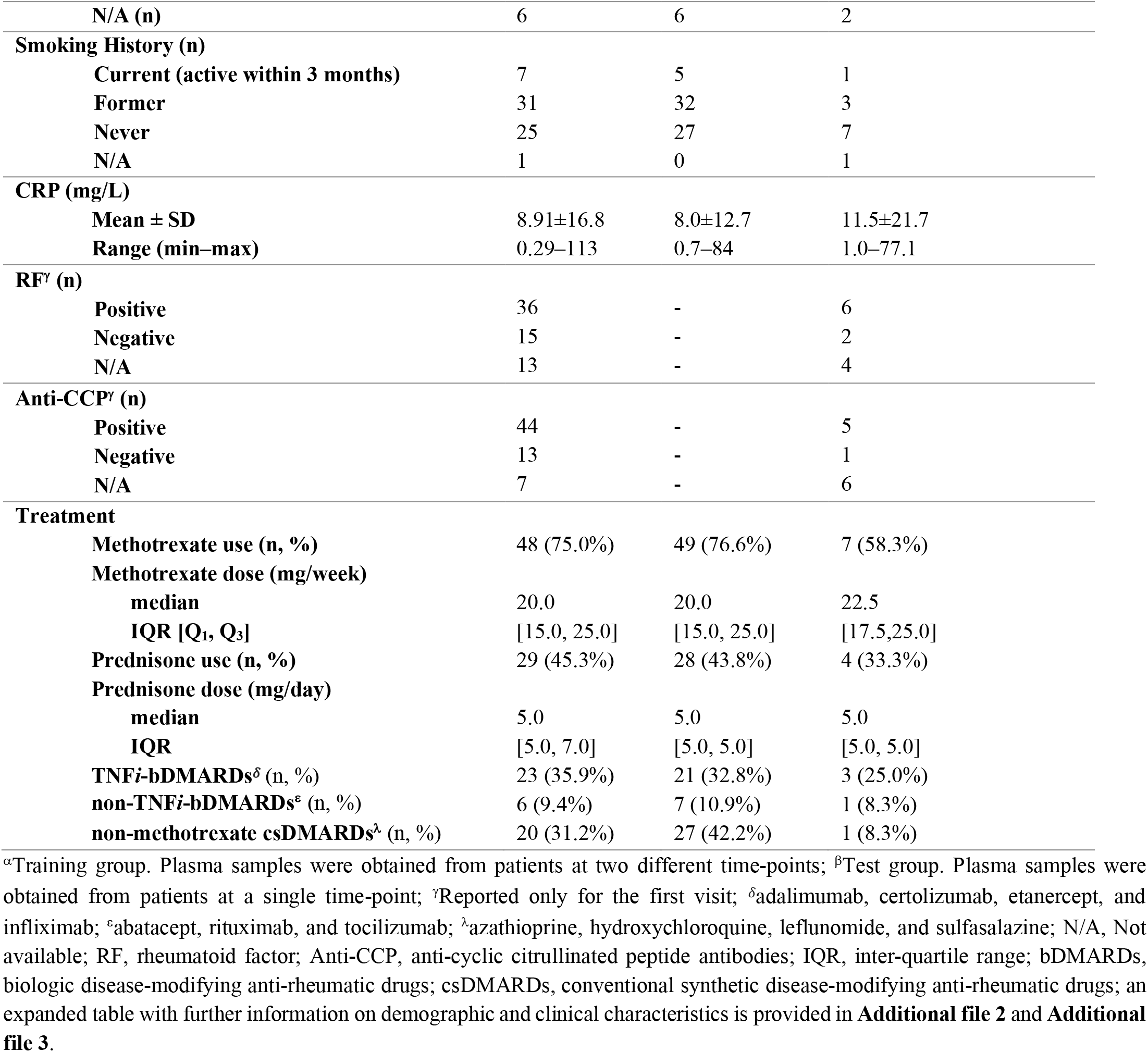
Demographic characteristics of study participants.

### Metabolomic Profiling

Untargeted metabolomic profiling of plasma samples from both discovery and validation cohorts through ultra-high performance liquid chromatography-tandem mass spectrometry (UPLC-MS/MS) was performed by Metabolon Inc. (Durham, NC, USA)’s Discovery HD4™ platform. Detailed descriptions of all methods regarding metabolomic profiling are available in **Additional file 4**.

### Analysis Workflow

Figure 1 provides a summary of the analytic strategy used on the 128 plasma samples of the discovery cohort to identify associations between metabolites and RA disease activity. The analysis workflow consists of two complementary approaches: Using mixed-effects logistic regression, the first approach identifies metabolites that are differentially abundant between higher and lower disease activity groups, which were determined by DAS28-CRP scores [18-20, 31] (**Fig. 1A**); the second approach uses mixed-effects linear regression to model the relationship between DAS28-CRP and metabolite abundances, allowing the detection of key biochemical features that associate with quantitative disease activity (**Fig. 1B**). To test the predictive accuracy of these selected features when incorporated into a generalized linear model, an additional cohort of twelve plasma metabolomic profiles (from twelve RA patients obtained at single time-points) was collected as an independent validation set.

### Pre-processing of Metabolomic Profiling Data

Statistical analyses on untargeted metabolomic data were performed using scaled imputed data provided by Metabolon, Inc. Briefly, the raw data were normalized to account for inter-day variation, which is a result of UPLC-MS/MS runs over multiple days, then the peak intensities were rescaled to set each metabolite’s median equal to 1. Missing values were then imputed with the minimum observed value of the metabolite across all samples, finally yielding the scaled imputed data. In addition, metabolites with missing values in over 20% of the entire samples were removed, resulting in 686 metabolites remaining for further analysis. R (v3.6.1), lme4 package (v1.1.21) [32], Python3 (v3.7.5), and sklearn (v0.22.2) were used to perform all data pre-processing and statistical analyses.

**Figure 1.**
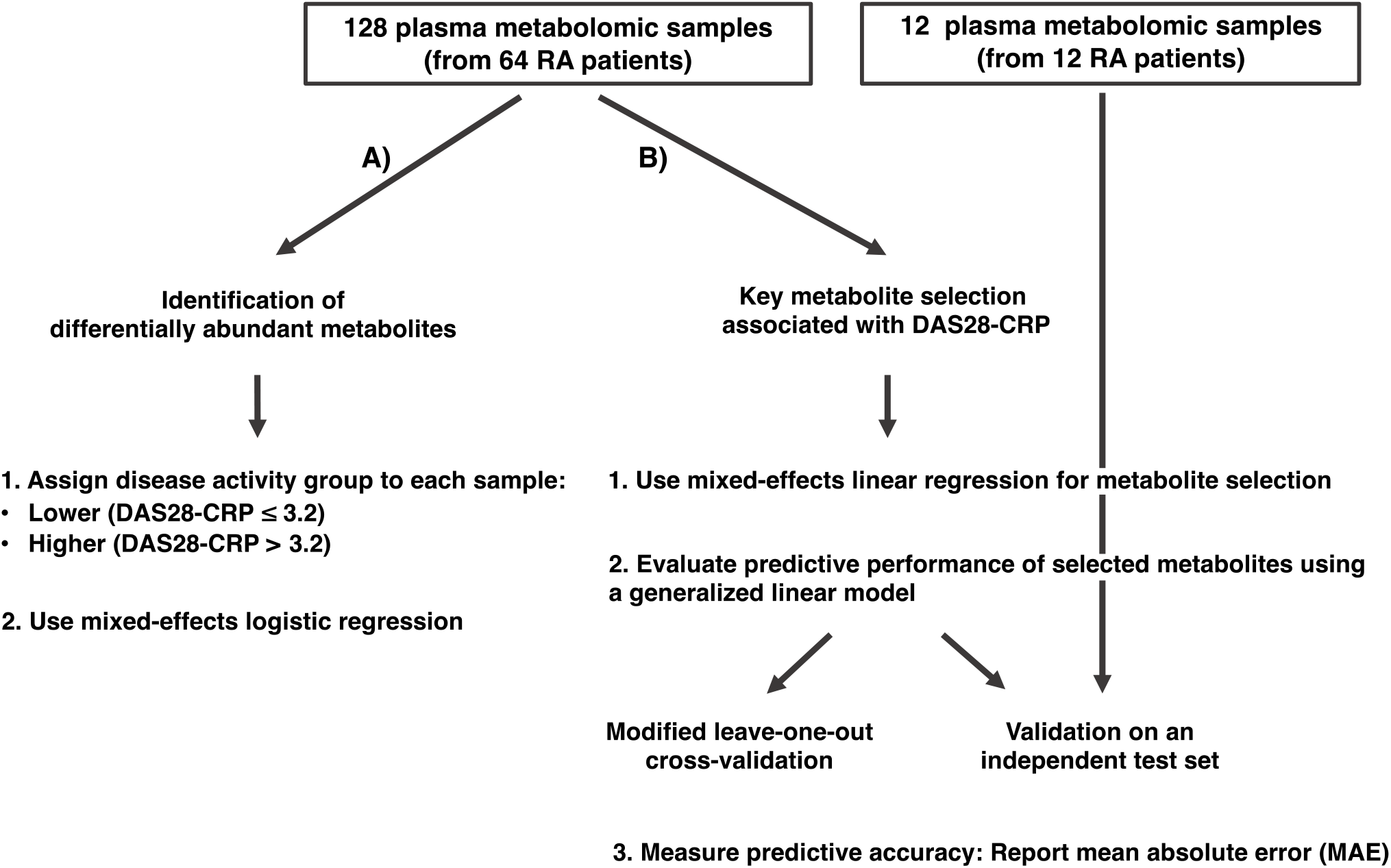
A multi-approach discovery strategy to identify metabolites indicative of RA disease activity. **(A)** Differentially abundant metabolites between higher and lower disease activity groups were identified using a mixed-effects logistic regression model adjusted for patient age and sex, as well as for Patient ID to control for having multiple samples from the same patient. **(B)** A selection scheme to identify metabolites associated with DAS28-CRP. Metabolites were selected with mixed-effects linear regression. To further demonstrate their association with DAS28-CRP, these metabolites were used to construct a generalized linear model for predicting DAS28-CRP. Predictive performance of the model was evaluated on the discovery cohort (using a cross-validation technique) and on a validation cohort.

### Delineation of RA Disease Activity Groups

Following previous reports [18-20, 31], samples from RA patients were divided into two disease activity groups based upon DAS28-CRP: ‘lower’ (DAS28-CRP ≤ 3.2, n = 76) and ‘higher’ (DAS28-CRP > 3.2, n = 52). These pre-defined two disease activity groups were used as the nominal response variable in a mixed-effects logistic regression model to identify differentially abundant metabolites between the two groups.

### Identification of Differentially Abundant Metabolites While Controlling for Confounding Factors

The following patient characteristics were examined to identify potential confounding factors in the association between plasma metabolites and disease activity (i.e., higher or lower disease activity) : age, sex, BMI, smoking history, and treatment use (for methotrexate, prednisone, non-methotrexate csDMARDs, TNF*i*-bDMARDs, and non-TNF*i*-bDMARDS). Based upon the Fisher’s exact test, patient age (age ≤ 60, age > 60) and sex (male, female) were observed to have statistically significant associations with the two disease activity groups; the *P*-value for age and sex was *P* = 0.01 (odds ratio [OR] = 2.74, 95% confidence interval [CI] = 1.15–6.73) and *P* = 0.02 (OR = 0.37, 95% CI = 0.14–0.88), respectively. On the other hand, no statistically significant associations were observed between these two disease activity groups and BMI (BMI ≤ 30, BMI > 30; *P* = 0.32), disease duration (duration ≤ 9 years, duration > 9 years; *P* = 0.14), smoking history (smoked at least once, never smoked; *P* = 0.36), or treatment use (user, non-user) for methotrexate (*P* = 0.83), prednisone (*P* = 0.58), TNF*i*-bDMARDs, i.e., adalimumab, certolizumab, etanercept, and infliximab (*P* = 0.18), non-TNF*i*-bDMARDs, i.e., abatacept, rituximab, and tocilizumab (*P* = 0.76), or other non-methotrexate csDMARDs, i.e., azathioprine, hydroxychloroquine, leflunomide, and sulfasalazine (*P* = 0.71). In addition, no significant changes in treatment use were observed between the two visits; *P*-values of the associations between treatment use and time-point based upon McNemar’s Chi-squared test for paired nominal data were as follows: methotrexate (*P* = 1), prednisone (*P* = 1), TNF*i*-bDMARDs (*P* = 0.75), non-TNF*i*-bDMARDs (*P* = 1), and non-methotrexate csDMARDs (*P* = 0.07). Therefore, the mixed-effects logistic regression model was adjusted for age and sex as fixed effects, but not for all other aforementioned covariates. In accordance with these results, age and sex have been previously reported to be connected to RA disease activity [33-35]. Herein, patient ID was considered as a random effect in the model to account for intra-subject variance due to having repeated measurements from a single patient. By controlling for patient ID (which are unique to each patient) as a random effect, we are acknowledging the non-independence in our data, that is, sampling that has taken place from within a patient. Leveraging multiple samples from the same patient allows us to compensate for the small number of samples in higher disease activity (DAS28-CRP > 3.2) in each visit (visit 1 and visit 2 having 25 and 27 samples, respectively) by maximizing the degree of freedom for the quantitative disease activity measure, and thereby to boost statistical power. Importantly, no significant difference was observed in DAS28-CRP between visit 1 and visit 2 (*P* = 0.98, Wilcoxon signed-rank test). Metabolites whose corresponding coefficients of the regression model were of *P*-value < 0.05 were considered as differentially abundant, that is, having a statistically significant association with disease activity group.

### Selection of Metabolites Associated with DAS28-CRP

Selection of metabolites associated with DAS28-CRP was performed with a mixed-effects linear regression model (DAS28-CRP as the continuous response variable), which controls for fixed effects (scaled metabolite abundances, patients’ age and sex) and for random effects (patient ID). Satterthwaite’s degrees of freedom method supported by lmerTest (v3.1.1) [36] was applied to test for the statistical significance (*P*-value) of associations between metabolites and DAS28-CRP. *P*-values were retrieved from the corresponding regression coefficients of the predictor variables.

### Evaluation of Predictive Performance of DAS28-CRP-associated Metabolites

A generalized linear model (GLM) was used to estimate DAS28-CRP scores using the aforementioned significantly associated metabolites as predictor variables. Predictive performance of the parameterized model was evaluated by two different techniques: First, a modified leave-one-out cross-validation approach was applied to the 128 samples of the training group (discovery cohort). More specifically, in each cross-validation loop, both samples from the same patient were allocated as an internal validation set, while all remaining samples (126 samples from 63 patients) were used to select metabolites significantly associated with DAS28-CRP (*P* < 0.05). These selected biochemical features were then included in a GLM for predicting DAS28-CRP scores of the remaining two samples (of the internal validation group) from their metabolite abundances. The second approach considers testing a GLM, which was composed of the DAS28-CRP-associated metabolites identified from all 128 samples of the training group, on the independent validation group of 12 plasma samples (validation cohort). For both techniques, model performance was reported using mean absolute error (MAE) and standard deviation (SD).

### Identification of Metabolites Associated with Treatment Use

A marginal, mixed-effects linear regression model was used to relate metabolite abundance with treatment use. Scaled metabolite abundance, treatment use, and patient ID was set as the response variable, predictor variable (fixed effect), and random effect, respectively. Use of the following treatments was assessed individually: methotrexate, prednisone, non-methotrexate csDMARDs, TNF*i*-bDMARDs, and non-TNF*i*-bDMARDs (names of individual drugs in each treatment group are provided in the footnote of **Table 1**). *P*-values were retrieved from the corresponding regression coefficient of the predictor variable (i.e., use or non-use), and a significance of *P* < 0.05 was reported as statistically significant.

### Identification of Differentially Abundant Metabolites Between Two CRP Groups

Metabolites that are significantly associated with disease activity groups and DAS28-CRP scores were further investigated to find those associated with patient groups delineated by CRP levels. First, all samples were divided into two groups as follows: ‘high-CRP’ (CRP *>* 3.0 mg/L, n = 52) and ‘low-CRP’ (CRP ≤ 3.0 mg/L, n = 76). Next, a marginal, mixed-effects linear regression model was used to define the abundance of a metabolite based upon the following fixed effects: CRP group, sex, age, smoking history, and treatment with prednisone, methotrexate, non-methotrexate csDMARDs, TNF*i*-bDMARDs; or non-TNF*i*-bDMARDs. Additionally, patient ID was treated as a random effect. Any covariates whose association with metabolite abundance was statistically significant (i.e., *P*-value of the corresponding regression coefficient < 0.05) were then included in an adjusted mixed model for metabolite abundance. Finally, metabolites were considered as differentially abundant between the two CRP groups if the association between metabolite abundance and CRP group was still found to be significant in the adjusted model (*P* < 0.05).

## Results

### Differentially Abundant Metabolites between Higher and Lower Disease Activity Groups

As shown in our analysis workflow (**Figure 1**), we first sought metabolites that were significantly different in abundance between two major disease activity groups. For this, we divided the 128 metabolomic profiles into two major categories (‘higher’ vs. ‘lower’) based upon the reported disease activity of the corresponding patient at the time of sample collection (**Materials and Methods**). Using a mixed-effects logistic regression model (**Materials and Methods**), we identified 33 metabolites as differentially abundant between higher (n = 52) and lower (n = 76) DAS28-CRP groups (**Fig. 2**). Most of these metabolites (31 of 33) were observed to have significantly increased abundances in lower disease activity, whereas the remaining two (glucuronate and hypoxanthine) were found to be significantly increased in higher disease activity. Notably, of the 31 metabolites increased in lower disease activity, seven metabolites (3-hydroxydecanoylcarnitine, dihomo-linoleoylcarnitine (C20:2), eicosenoylcarnitine (C20:1), linoleoylcarnitine (C18:3), linoleoylcarnitine (C18:2), stearoylcarnitine (C18), palmitoylcarnitine (C16)) are a part of acylcarnitine metabolism, and represent a 3.6-fold enrichment in metabolites involved in this particular pathway (*P* = 1.9 × 10^−3^, hypergeometric test). It is important to note that the differences seen are relatively small in terms of fold-change, with most of the metabolites varying by 1.1 –1.3 fold. Despite these subtle differences within RA patients of varying disease activities, we were still able to obtain statistically significant signal even after considering and controlling for all known potentially confounding factors (which often leads to reduction in statistical power), while adhering to our cut-offs for statistical significance (*P* < 0.05).

**Figure 2.**
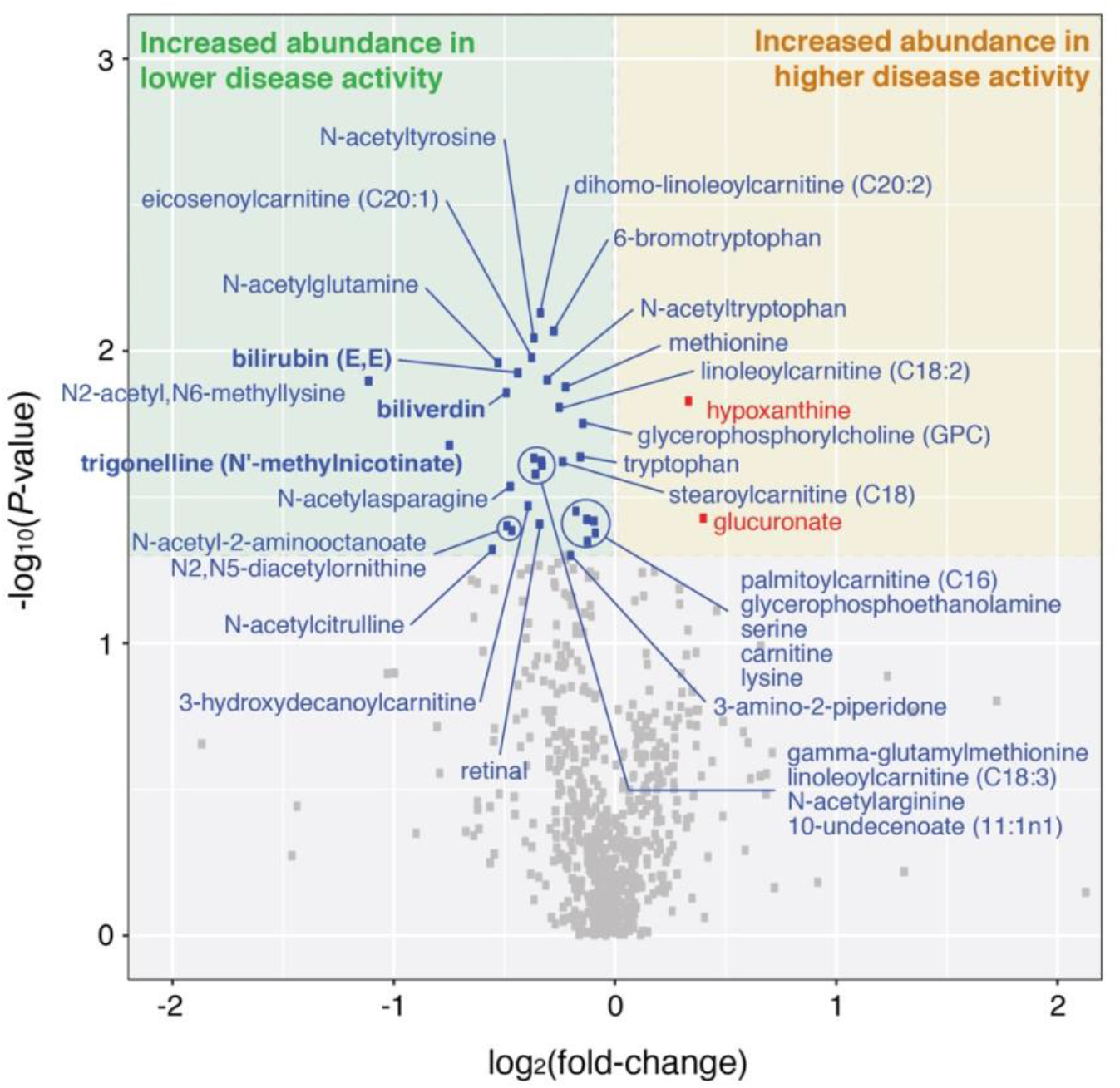
Plasma metabolites differentiating between higher and lower disease activity groups in RA. A total of 2 and 31 metabolites were found to be significantly increased in higher (DAS28-CRP **>** 3.2, n = 52) and lower (DAS28-CRP **≤** 3.2, n = 76) disease activity groups, respectively. Each point corresponds to a metabolite (686 total). Differentially abundant metabolites were found using a mixed-effects logistic regression model on the discovery cohort (128 samples), for which age and sex were adjusted. Metabolites with *P*-value < 0.05 (based upon the corresponding coefficient of the regression model) were considered as significantly different between groups. *P*-values and fold-changes for all metabolites are listed in **Additional file 5**. Metabolites in bold have been previously described in the literature for their associations with RA.

N2-acetyl,N6-methyllysine (|log_2_(FC)| = 1.11, *P* = 1.26 × 10^−2^) and trigonelline (N’-methylnicotinate) (|log_2_(FC)| = 0.74, *P* = 2.09 × 10^−2^), which were both found to have increased abundance in lower disease activity, were the top two metabolites having the largest fold-changes between the two groups. Although the direct relevance of N2-acetyl,N6-methyllysine to RA is currently not well understood, N2-acetyl,N6-methyllysine is part of the lysine metabolism pathway, which has been reported to be associated with RA in the following studies: i) according to Teitsma *et al*., serum metabolites associated with lysine degradation were observed to have a higher concentration in early RA patients who achieved sustained drug-free remission (after Tocilizumab-or Methotrexate-based treatment) compared to those who never achieved a drug-free status [24]; and ii) Yang *et al*. reported that metabolic products of lysine degradation (carnitine and pipecolic acid) were significantly increased in RA patients than in normal subjects [37].

In regard to trigonelline, which is a product of niacin (vitamin B3) metabolism, this alkaloid has been suggested to have therapeutic potential for diabetes and central nervous system disease [38], and also reported to demonstrate anti-inflammatory properties in mice [39]. In accordance with our results showing decreased abundance of trigonelline in higher disease activity, trigonelline could be of interest in future studies on inflammatory responses in RA.

Biliverdin (|log_2_(FC)| = 0.48, *P* = 1.38 × 10^−2^) and bilirubin (E,E) (|log _2_(FC)| = 0.43, *P* = 1.18 × 10^−2^), which are known metabolic products of the heme catabolic pathway, were also observed to have significantly increased abundances in lower disease activity. In particular, biliverdin has been shown to: i) inhibit the activation of pro-inflammatory transcription factors, including NFkB both *in vitro* and *in vivo* [40-44]; ii) inhibit the proliferation of primary T cells stimulated with anti-CD3 and anti-CD28 monoclonal antibodies by inhibiting NFAT/NF-kB activation in a mouse model of heart transplantation [45]; and iii) improve corneal inflammation mediated by heme-oxygenase 2 (HO-2) deficiency in a transgenic mouse model [41]. Moreover, bilirubin, which is derived from the reduction of biliverdin by biliverdin reductase, has been reported as a potential biomarker for RA in line with our findings. For example, Peng *et al*. observed a decreased concentration of serum bilirubin in RA patients compared to healthy controls, as well as in RA patients with worsening disease activity [46]. Additionally, Fischman *et al*. found that total bilirubin levels are inversely related to RA disease activity even after adjusting for multiple confounders (e.g., age, sex, race), and discussed the possibility of bilirubin (a known anti-oxidant) having a physiological anti-inflammatory effect [47]. This point is further elaborated upon by Jangi *et al* [48], who have described in detail the immunosuppressive properties of unconjugated bilirubin in RA and other inflammatory disorders. The full list of differentially abundant metabolites and their associated pathways are shown in **Additional file 5**.

### Metabolic Feature Selection Improves DAS28-CRP Prediction Accuracy

Having uncovered metabolites demonstrating altered abundance between two major disease activity groups, we next asked whether quantitative disease activity can be predicted with plasma metabolomes. As untargeted metabolomic profiling can yield a considerable amount of noise and random fluctuations in observed signal [49], it is necessary to first select informative metabolic features that reliably capture relevant aspects of the phenotype of interest [50]. For this, we used mixed-effects linear regression models to select metabolites significantly associated with DAS28-CRP. Afterwards, the abundances of the selected metabolic features were incorporated into a generalized linear model (GLM) to predict DAS28-CRP. For comparison purposes, a GLM was constructed without metabolic feature selection, and thereby taking into consideration all features of a metabolomic profile. Details regarding GLM construction and performance evaluation are provided in **Materials and Methods**.

When applying a modified leave-one-out cross-validation technique to the training group samples (n = 128), we found that the GLM incorporating metabolites that were significantly associated with DAS28-CRP outperformed the model without feature selection (i.e., using all metabolites). As shown in **Figure 3**, the distribution of absolute errors between the observed and predicted DAS28-CRP scores was smaller (with respect to the cumulative area under the error curve) for the GLM with feature selection than that without feature selection. To this point, the prediction MAE (±SD) of the GLM with and without feature selection was 1.51 (±1.89) and 2.02 (±2.52), respectively.

**Figure 3.**
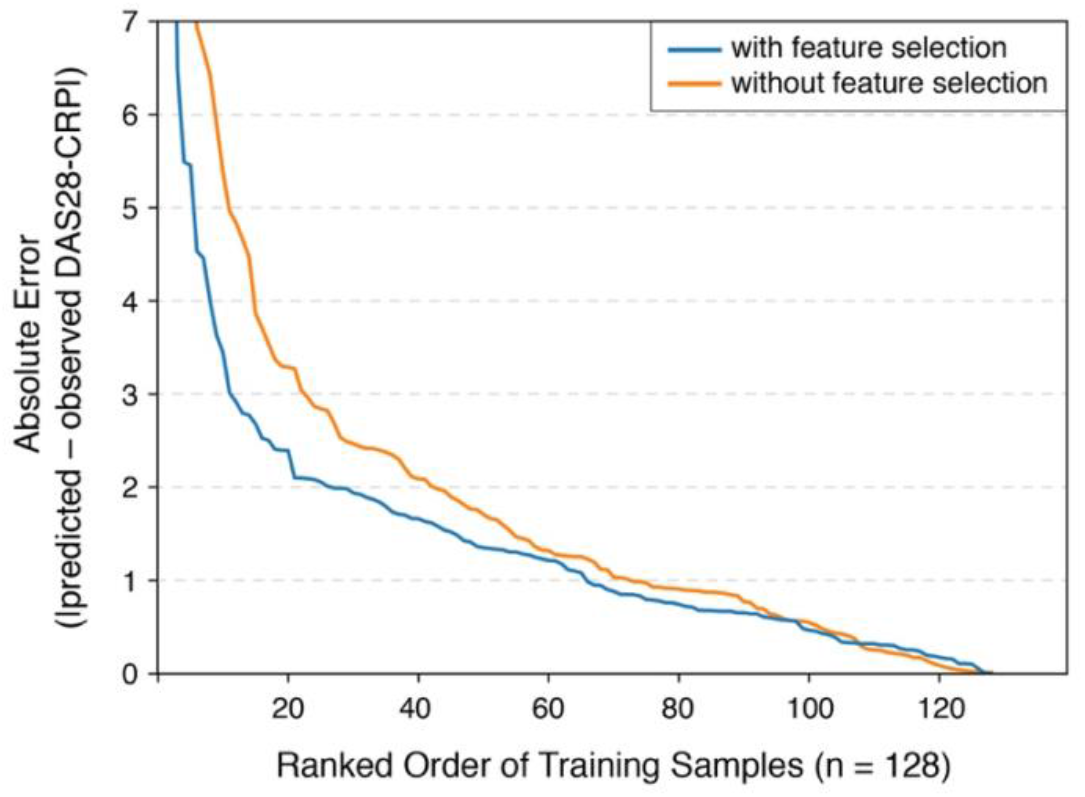
Evaluation of DAS28-CRP predictive performance in cross-validation. A modified leave-one-out cross-validation approach was used on the samples of the training group (128 samples) to test the performance of a generalized linear model (GLM) in predicting DAS28-CRP scores from metabolite abundances. Distributions of absolute errors from models with and without a feature selection scheme were compared to identify the more robust model. The GLM with the feature selection scheme performed better (MAE±SD: 1.51±1.89) than the model without feature selection (MAE±SD: 2.02±2.52).

Having confirmed that feature selection can lead to a more accurate prediction model in cross-validation, we applied the same scheme to all metabolome samples of the discovery cohort to obtain a final set of metabolites associated with DAS28-CRP (*P* < 0.05). After adjusting for potential confounding factors (**Materials and Methods**), this resulted in a collection of 51 plasma metabolites (**Table 2**). These metabolites were used to construct a final GLM, whose predictive accuracy was tested on an independent validation cohort (n = 12) of plasma metabolomic profiles from twelve RA patients (importantly, this additional cohort was not drawn from the same population distribution from which the features were derived). On this previously unseen cohort, the GLM constructed with only the 51 selected metabolites performed considerably better than the model without the feature selection scheme by over two-fold (**Fig. 4A**); the prediction MAE of the GLM with and without feature selection was 0.97 (±0.47) and 2.01 (±2.18), respectively. Likewise, when the actual and predicted DAS28-CRPs were plotted together for both GLMs (**Fig. 4B**), we found that the model with the selection scheme performed more favorably. More specifically, a stronger correlation between the actual and predicted disease activity scores was observed in the model with feature selection (Spearman’s *ρ* = 0.69, *P* = 1.40 × 10^−2^, 95% CI: [0.18, 0.90]) compared to the model without (Spearman’s *ρ* = 0.18, *P* = 5.72 × 10^−2^, 95% CI: [-0.44, 0.68]).

**Table 2.**
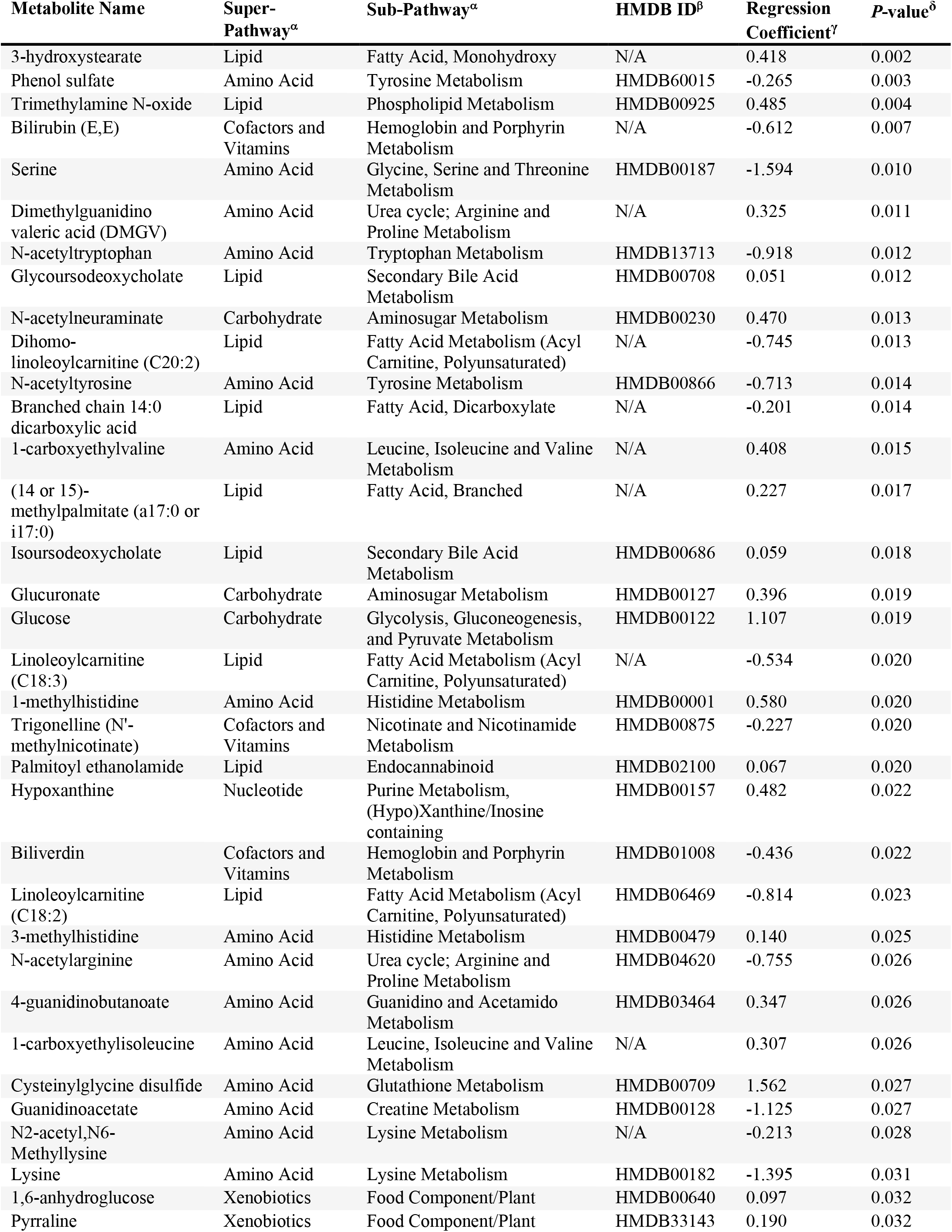
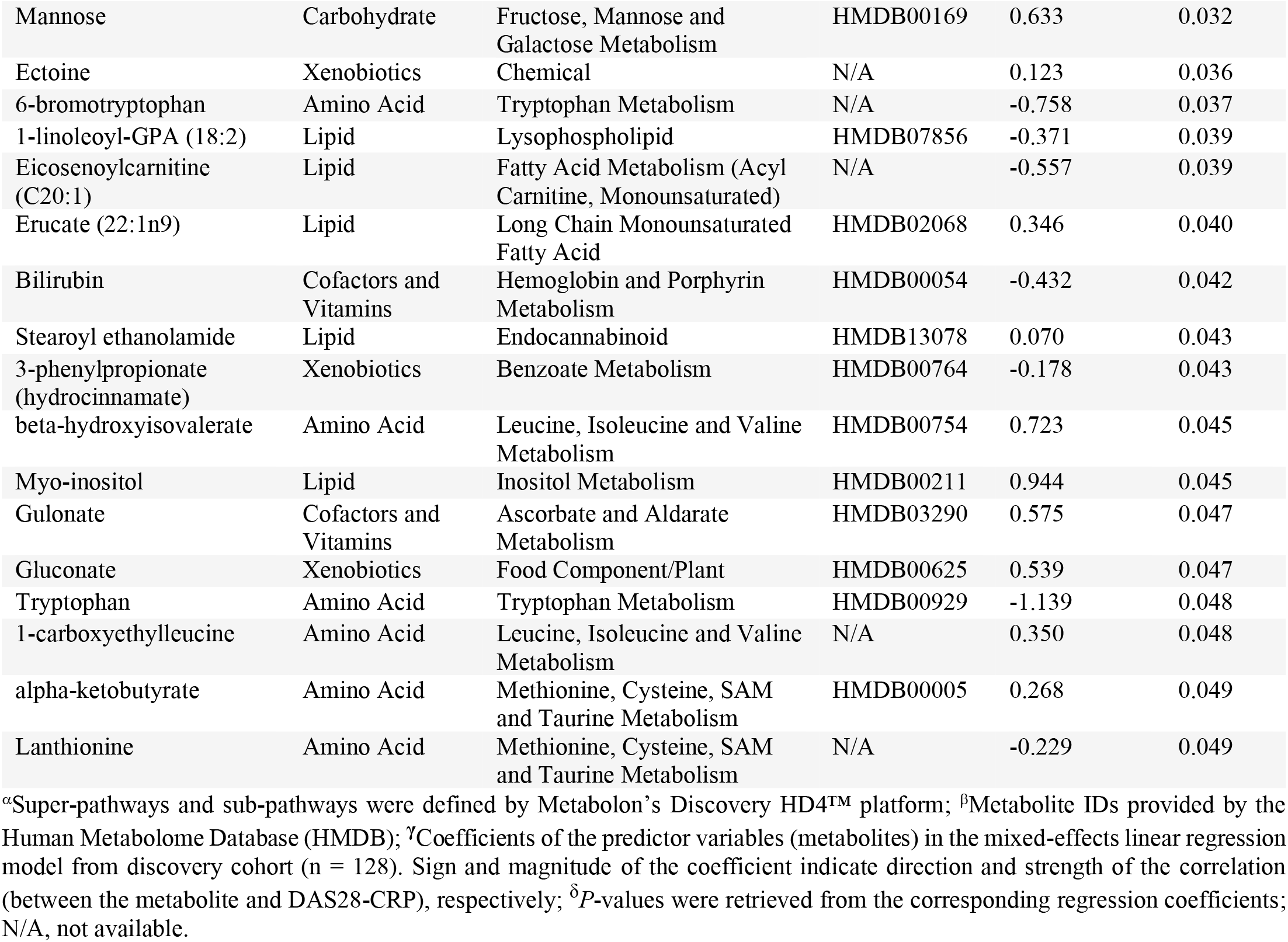
Plasma metabolites significantly associated with DAS28-CRP.

**Figure 4.**
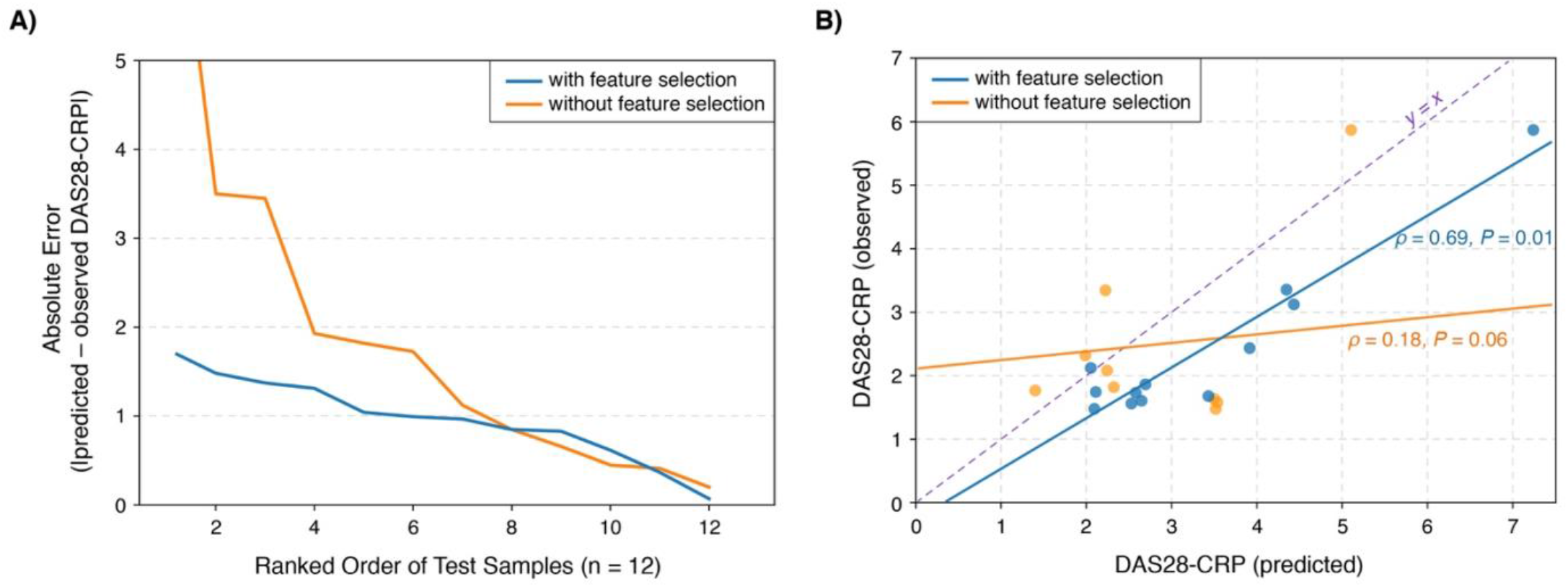
GLM with feature selection provides improved DAS28-CRP prediction accuracy in an independent validation group (12 samples). **(A)** Performance of GLMs in predicting quantitative disease activity were evaluated on samples of an independent validation group. Distributions of absolute errors from models with and without a feature selection scheme were compared to identify the more robust model. **(B)** Selection of metabolic features prior to model training resulted in higher predictive performance, as evidenced by the stronger correlation between observed and predicted DAS28-CRPs. Three samples predicted to have negative DAS28-CRP values are omitted from the scatter-plot. Dashed violet line indicates ‘y = x’, i.e., an exact match between the observed and predicted values. 95% confidence interval for *ρ* with feature selection: [0.18, 0.90]; without feature selection: [-0.44, 0.68].

### Commonly Identified Metabolites from Two Different Analytic Approaches

To summarize the findings above, we found that, from the 686 total detectable metabolites in a metabolomic profile, 33 (4.8%) were differentially abundant between higher and lower disease activity; and 51 (7.4%) were significantly associated with DAS28-CRP (**Fig. 5**). These separate findings amounted to a total of 67 unique metabolites, among which 40 were found to have no association with the use of prednisone, methotrexate, other non-methotrexate csDMARDs, TNF*i*-bDMARDs, or non-TNF*i*-bDMARDs (**Materials and Methods**). Notably, eight metabolites (6-bromotryptophan, bilirubin (E,E), biliverdin, glucuronate, N-acetyltryptophan, N-acetyltyrosine, serine, and trigonelline) were not only consistently detected across both analytic approaches, but also found to have no association with any treatment use; these results strongly suggest key metabolic pathways and modules potentially contributing to, or serving as indicators of, RA pathogenesis independent of confounding treatment effects. Consistent with this idea, additional studies into the metabolites found in this study (the majority of which have yet to be linked to RA) may be able to provide new insight into the perturbed physiological metabolic processes—which are then in turn reflected in blood—underlying disease progression in RA.

**Figure 5.**
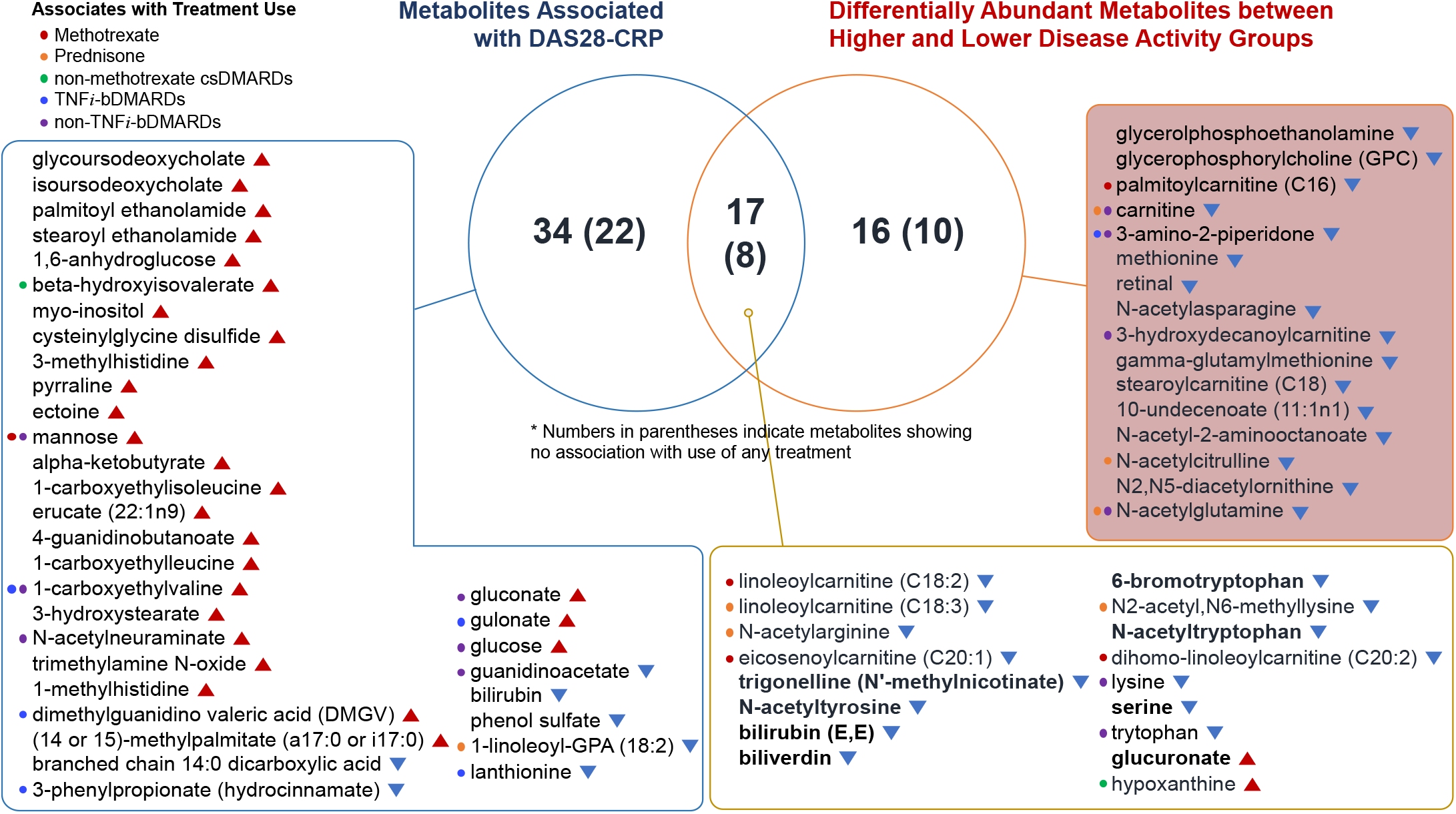
Venn diagram of all plasma metabolites identified through the multi-approach discovery strategy. A total of 67 unique metabolites were identified, among which 40 were found to have no association with the use of treatment. Notably, eight metabolites (6-bromotryptophan, bilirubin (E,E), biliverdin, glucuronate, N-acetyltryptophan, N-acetyltyrosine, serine, and trigonelline) in bold were not only consistently detected across both analytic approaches, but also found to have no association with any treatment use. Colored circles indicate metabolites whose abundances associate with treatment use. Metabolites with red triangles were found to have increasing abundances with worsening disease activity, whereas metabolites with blue triangles were found to have decreasing abundances with worsening disease activity.

### Metabolites Associated with CRP Patient Groups

Elevated levels of C-reactive protein (CRP) in the blood is well known to often indicate increased inflammatory conditions, which may be caused by a wide variety of acute (e.g., infections) and chronic disorders (e.g., rheumatoid arthritis, inflammatory bowel disease). In RA patients, CRP levels have been observed to increase after acute mental stress tasks [51], and also to be linked to risk of cardiovascular disease [52]. Furthermore, several serum metabolites were found to reflect inflammatory activity in patients with early arthritis [53].

We further investigated the aforementioned 67 plasma metabolites to see whether any were differentially abundant between two CRP patient groups, i.e., ‘high-CRP’ (CRP *>* 3.0 mg/L, n = 52) and ‘low-CRP’ (CRP ≤ 3.0 mg/L, n = 76) (**Materials and Methods**). While controlling for potential confounding variables, we identified eight total metabolites that were significantly associated with CRP patient group. More specifically, the abundances of mannose, beta-hydroxyisovalerate, (14 or 15)-methylpalmitate (a17:0 or i17:0), erucate (22:1n9), 10-undecenoate (11:1n1), N-acetylcitrulline were higher in high-CRP, while those of serine and linoleoylcarnitine (C18:3) were lower in high-CRP (**Fig. 6**). Application of these plasma metabolites, which were found to be connected to both RA disease activity and circulating CRP levels, may lead to the development of new clinical laboratory tests to further enable precision medicine for RA patients.

**Figure 6.**
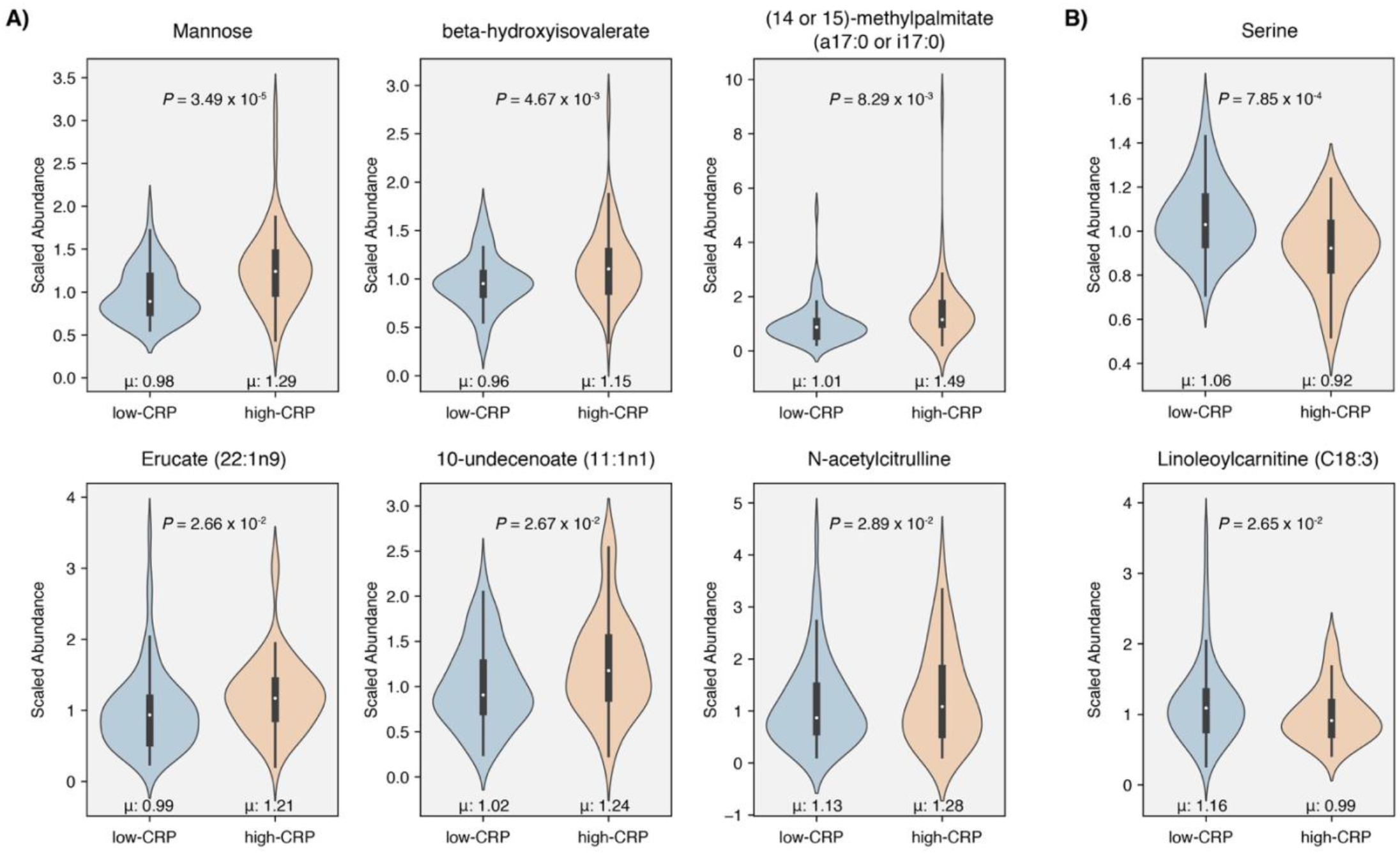
Metabolites differentially abundant between two CRP patient groups. Among the 67 total metabolites identified through our multi-approach analysis on the discovery cohort (n = 128), eight metabolites were identified to have significant associations with CRP group while controlling for confounding variables (regression coefficient for CRP, *P* < 0.05) **(A)** Metabolites with higher abundances in the high-CRP group: mannose, beta-hydroxyisovalerate, (14 or 15)-methylpalmitate (a17:0 or i17:0), erucate (22:1n9), 10-undecenoate (11:1n1), and N-acetylcitrulline. **(B)** Metabolites with higher abundances in the low-CRP group: serine and linoleoylcarnitine (C18:3).

### Plasma metabolites associate with clinical improvement in RA

Based upon the European League Against Rheumatism (EULAR) response criteria for DAS28-CRP [54], we found that sixteen of the 64 patients in the discovery cohort showed moderate or good improvement in disease activity from visit 1 to visit 2, while the remaining 48 patients did not show clinical improvement at the time of their second visit. This discovery provided an entry point for the following analysis: For each of these two patient groups, i.e., ‘Improved’ (n = 16) and ‘Non-improved’ (n = 48) patients, we aimed to identify metabolites whose abundances significantly changed from visit 1 to visit 2, while controlling for the same confounding factors (mixed-effects regression model, *P* < 0.05). As a result, we identified eleven metabolites whose abundances significantly changed in the Improved patient group (**Additional file 6)**, while nineteen metabolites showed significant changes in the Non-improved patient group (**Additional file 7)**. The following three metabolites, which were discovered in our previous analyses on the 128 plasma metabolome samples of the discovery cohort, were detected once again: erucate (22:1n9), a metabolite identified to be associated with both DAS28-CRP and CRP patient group, was identified to be significantly different between visit 1 and visit 2 in patients who did not show clinical improvement (Non-improved); 3-amino-2-piperidone, a metabolite identified to be differentially abundant between higher and lower disease activity in our study, was identified to be significantly different between visit 1 and visit 2 in patients who showed clinical improvement (Improved); and gamma-glutamylmethionine, a metabolite identified to be differentially abundant between higher and lower disease activity, was identified to be significantly different between visit 1 and visit 2 in the Non-improved group. These results allow us to expand our future direction to investigate metabolites associated with clinical improvement in patients with RA.

## Discussion

Dysfunctions in cellular and tissue metabolism are involved in a broad range of autoimmune disorders [55-58], including RA [59-61]. These metabolic implications highlight the importance of investigating which biochemical functions and metabolic states are altered during the onset and progression of disease. To this end, metabolomics platforms (and the accompanying wealth of data) can present unique opportunities for discovering novel disease ‘molecular signatures’ [62], which can be interpreted through the lens of annotated biochemical relationships. Moreover, high-throughput profiling can enable the identification of circulating pro-inflammatory (disease-triggering) and anti-inflammatory (disease-protective) metabolites in RA, as elaborated upon by Coras *et al*. [63]; such discoveries may facilitate the design of either dietary or gut microbiome-based intervention strategies to improve wellness or alter the course of disease for RA patients. In this study, by performing a global metabolomic profiling analysis on 128 plasma samples obtained from 64 patients with RA, we identified biochemical signatures associated with, and predictive of, disease activity. Mainly, through a combination of statistical approaches for metabolic signature discovery, we identified several metabolites that: i) differ significantly between lower and higher disease activity groups; and ii) significantly associate with DAS28-CRP. Of note, our study is the first to leverage biochemical features from a plasma metabolomic profile to predict quantitative disease activity.

Interestingly, we identified eight metabolites (6-bromotryptophan, bilirubin (E,E), biliverdin, glucuronate, N-acetyltryptophan, N-acetyltyrosine, serine, and trigonelline) that were commonly found across different statistical approaches, possibly capturing representative metabolite signals of RA progression. We discussed above the reported roles of bilirubin and biliverdin in RA. Moreover, these two metabolites were previously reported for their cytoprotective and anti-inflammatory effects [64-68], and even suggested as an “RA protective factor” by Fischman *et al*. [47]. Interestingly, high concentrations of bilirubin and biliverdin were reported in other inflammatory disorders, such as atherosclerotic diseases [69] and autoimmune encephalitis [67]. In regards to the other six metabolites, clear and definitive connections with RA have not yet been established. However, if our results on bilirubin and biliverdin were to serve as benchmarks for reliably identifying plasma metabolites important to RA disease activity, then these remaining metabolites may be deemed as leading candidates for future investigations.

Glucuronate was found to show elevated abundance in higher disease activity than in lower disease activity. This glucose-derivative is involved in the detoxification of xenobiotics via glucuronidation in the mammalian liver. Interestingly, this process can be reversed by gut bacteria harboring β-D-glucuronidases [70], and thereby releasing (potentially toxic) exogenous compounds into the gut lumen and subsequently into circulation [71, 72]. In that respect, examining a possible role involving dysbiosis in the gut microbiota— combined with metabolomic approaches to infer relationships between gut microbes and blood metabolites in RA, as shown by Chen *et al*. [73]—may help elucidate a microbial-based mechanism explaining the observed alterations in plasma glucuronate.

Serine was seen to decrease with worsening disease activity. In line with our results, albeit in an RA mouse model with collagen-induced arthritis (CIA), plasma levels of serine and other free amino acids were found to have significantly decreased in the CIA group compared to control mice [74]. In another study wherein synovial fluid of RA patients were examined for citrullinated proteins (which is widely known to result in a rise in anti-citrullinated protein antibodies in RA), Tilvawala *et al*. found increased citrullination in a wide array of serine protease inhibitors (Serpins) and serine proteases [75]; in the same study, the investigators demonstrated *in vitro* that citrullinating serine protease inhibitors nearly abolishes their inhibitory activity towards their target proteases. Although we have yet to uncover whether a decrease in plasma serine levels (with worsening disease activity) is linked to citrullination of serine proteases in synovial fluid, we speculate that changes in serine may reflect dysregulated protein degradation during systemic inflammatory activity and joint destruction in RA.

We note a few limitations of this study: First, we acknowledge that our study includes a relatively small number of samples within each disease activity group of the discovery cohort and the validation cohort. Nevertheless, we were able to detect statistically significant metabolites in all analytical strategies, demonstrating that our data provides reasonably sufficient statistical power. Certainly, a much larger cohort would have been ideal; however, this is a small pilot study on our stored plasma samples, and obtaining an additional cohort is outside of its scope. Encouragingly, despite the low sample size of the validation cohort, we were able to successfully show that feature selection is a necessary step in the model-training process, and we expect this finding to translate well to larger cohorts in our future studies. Nevertheless, in order to more meticulously examine the role of blood metabolites in RA, future investigations will warrant a higher number of samples and more detailed subject characteristics. Second, to define RA disease activity, we solely used the DAS28-CRP scoring system, which is dependent upon acute-phase responses that may not accurately reflect patients who have an inflammation-free state [24]. Our future plans include performing our analytical pipeline with other RA disease activity metrics (e.g., clinical disease activity index (CDAI), simple disease activity index (SDAI)) to test the robustness of our findings. Third, all of our multivariate analyses followed adjustment for patient age and sex only. Other potential confounders that may affect the concentration of blood metabolites, such as diet, exercise habits, lifestyle factors, time of the day of sample collection, and gut microbiome, were not considered as predictor variables in our analyses. Fourth, comorbidity can certainly be a significant confounding factor. Alternatively, comorbidities could theoretically be contributors or mediators to inflammatory disease activity in patients with RA. At this stage, it would have been premature to adjust statistically for the effects of particular comorbidities or for the presence of multi-morbidity (i.e., multiple chronic conditions) before carefully investigating for their potential interaction with p lasma metabolites and RA disease activity. This may lead to ‘over-adjustment’ and falsely concluding that certain metabolites are not significant when they may, in fact, be very important. Future studies will be necessary to explore potential interactions between comorbidities, RA disease activity, and plasma metabolite levels in RA. Last, despite the similarities in our findings with previous investigations (as noted above), many of our results are reported for the first time and remain to be validated by others. Possible causes of discrepancies with the work of others include the comparatively small number of samples in this pilot study; technical and biological sources of random noise; the uniqueness of our recruited patient cohort; variabilities in detection protocols and instrument sensitivity; and use of alternate statistical techniques and potential over-fitting. Future efforts, by us and others, are likely to elucidate truly robust signal and collectively strengthen the confidence in our novel findings.

Despite the aforementioned limitations, our study establishes the far-reaching utility of using cutting-edge technological and analytical approaches for plasma metabolomic profiling and justifies analogous investigations at larger scales. The identified metabolites could be a reflection of the perturbed metabolic processes concurrent with worsening disease activity, and our findings will inspire future studies into how inflammation and pain in RA are coupled to physiological metabolism. Moreover, our identified sets of signature metabolites offer a promising glimpse into biomolecular marker panels for diagnosing disease activity of RA patient s solely through blood (thereby complementing current diagnostic approaches), with the overall aim to make such assessments faster, cheaper, and less invasive. In turn, studies such as ours are expected to contribute towards fully realizing the potential of virtual and digital healthcare by foregoing the need for patients to physically arrive at the clinic to meet their primary care provider in person.

As the gut microbiome has been recognized to be implicated in RA [73, 76-78]—possibly through complex mechanisms underlying microbe-microbe and host-microbe biochemical cross-talk [79]—integrated profiling of both stool metagenome and blood metabolome would provide an in-depth, comprehensive view of functional dysbiosis during RA onset and progression. Interestingly, a recent study showed that blood metabolites can be predictive of gut microbiome alpha-diversity [80]. Such investigations into integrating across multiple data types of the same phenotype can help to amplify the primary biological signal of interest relative to noise, as well as provide actionable insights. In conclusion, the results reported herein are poised to eventually improve disease management and outcomes of patients with RA and other rheumatic diseases, as well as to provide novel means of monitoring health and wellness [81].

## Data Availability

Raw metabolomic datasets, as well as source codes used to reproduce the results in this study, are available at https://github.com/jaeyunsung/RA_plasma_metabolomics_2020.

## List of Abbreviations

RA: Rheumatoid arthritis
DMARDs: Disease-modifying anti-rheumatic drugs
bDMARDs: Biologic disease-modifying anti-rheumatic drugs
csDMARDs: Conventional synthetic disease-modifying anti-rheumatic drugs
UPLC-MS/MS: Ultra-high performance liquid chromatography-tandem mass spectrometry
DAS28-CRP: Disease Activity Score-28 using C-reactive protein
BMI: Body mass index
GLM: Generalized linear model
MAE: Mean absolute error
SD: Standard deviation
ESR: Erythrocyte sedimentation rate
CRP: C-reactive protein
Anti-CCP: anti-cyclic citrullinated peptide antibodies
HMDB: Human metabolome database
CIA: Collagen-induced arthritis
CDAI: Clinical disease activity index
SDAI: Simple disease activity index

## Supplementary Information

Additional file 1: Supplementary Figure 1, Histogram of DAS28-CRPs corresponding to the 128 total samples of the discovery cohort.

Additional file 2: Supplementary Table 1, Subject characteristics of 64 patients (128 samples) from the discovery cohort.

Additional file 3: Supplementary Table 2, Subject characteristics of 12 patients (12 samples) from the validation cohort.

Additional file 4: Supplementary Information on detailed methods regarding metabolomic profiling (Metabolon).

Additional file 5: Supplementary Table 3, Differentially abundant metabolites between lower (DAS28-CRP ≤ 3.2) and higher (DAS28-CRP > 3.2) disease activity groups.

Additional file 6: Supplementary Table 4, Metabolites displaying significant changes in abundances in patients with clinical improvement.

Additional file 7: Supplementary Table 5, Metabolites displaying significant changes in abundances in patients without clinical improvement.

## Declarations

### Ethics approval and consent to participate

This study was approved by the Mayo Clinic Institutional Review Board (no. 14-000616 and no. 14-000680) in accordance with the Declaration of Helsinki. All methods and procedures were performed in accordance with Mayo Clinic Institutional Review Board guidelines and regulations.

### Consent for publication

All patients provided written informed consent.

### Availability of data and materials

All raw metabolomic datasets, as well as source codes used to reproduce all results in this study, are available at https://github.com/jaeyunsung/RA_plasma_metabolomics_2020.

### Authors’ contributions

B.H., J.M.D., and J.S. conceived the problem and designed all analytical methodologies. B.H. performed all computational analyses. All authors analyzed the data. B.H., J.M.D., and J.S. wrote the manuscript, with editorial contributions from other authors. J.M.D. is the principal investigator of the Mayo Clinic Rheumatology Biobank, from which blood samples were collected from patients with rheumatoid arthritis. All authors reviewed and approved the final manuscript.

## Acknowledgements

First and foremost, we thank our dear patients who volunteered for this study. We also thank the Mayo Clinic Division of Rheumatology study coordinators (Jennifer Sletten and Kathleen McCarthy-Fruin) for their help in making this work possible.

